# Kidney Tubule Dysfunction and Injury and the Long-Term Risk of Acute Kidney Injury Following Cardiac Artery Bypass Graft Surgery

**DOI:** 10.1101/2025.08.26.25334515

**Authors:** Lauren Shingler, Ashutosh Tamhane, Ching-Min Chu, Jennifer A. Frey, Emily B. Levitan, Suzanne E. Judd, Alexander L. Bullen, Edward D. Siew, Joseph V. Bonventre, Michael G. Shlipak, Byron Jaeger, Jesse Seegmiller, Alexander Keister, Orlando M. Gutierrez, Joachim H. Ix, Henry E. Wang

**Author notes:** Corresponding Author: Henry E. Wang (HW).

## Abstract

**Objective:** Determine whether urine biomarkers of kidney tubule dysfunction and injury are associated with future-term risk of post-CABG AKI.

**Design:** Nested cohort study using data from the REasons for Geographic And Racial Differences in Stroke (REGARDS) study.

**Setting:** A national, population-based, longitudinal cohort study of 30,239 U.S. adults aged ≥45 years.

**Participants:** We included all 760 participants who underwent CABG surgery. The final cohort included 394 participants after exclusion for a history of dialysis or kidney transplant at admission (n=3), illegible chart data (n=346), missing laboratory data (n=15) or underwent a cardiac procedure mislabeled as CABG (n=2).

**Exposures:** Using spot urine obtained at enrollment, an average of 5.5 years before CABG surgery, we measured biomarkers of kidney tubule dysfunction (urine alpha-1 microglobulin [A1M], uromodulin [UMOD] and epidermal growth factor [EGF]) and injury (kidney injury molecule-1 [KIM-1]).

**Main Outcomes:** AKI development following surgery, defined as an increase in serum creatinine ≥0.3 mg/dL from 48 hours prior to CABG to end of hospitalization.

**Results:** Of 394 eligible participants who underwent CABG (mean age 66, 29% female, 20% Black), 176 (45%) experienced post-operative AKI. Higher baseline urine A1M was associated with higher odds of AKI (adjusted OR 1.34 per 2-fold higher A1M, 95% Confidence Interval (CI): 1.00–1.80). Higher urine UMOD was associated with lower odds of AKI (adjusted OR 0.77 per 2-fold higher UMOD, 95% CI 0.62–0.95). Higher EGF showed a non-significant tendency towards lower odds of AKI (adjusted OR 0.79 per 2-fold higher EGF, 95% CI 0.59-1.05). KIM-1 was not associated with AKI (adjusted OR 0.92 per 2-fold higher KIM-1, 95% CI 0.77– 1.10).

**Conclusions:** Select biomarkers of tubule dysfunction, but not injury, measured when patients were at a stable baseline, are associated with future AKI after CABG. These markers of kidney dysfunction may offer a strategy for identifying individuals vulnerable to AKI after CABG.

## Introduction

Acute kidney injury (AKI) is a significant burden on the healthcare system accounting for $10 billion per year in additional healthcare costs.[1] In patients undergoing coronary artery bypass graft (CABG) surgery, approximately one in five will develop AKI post-operatively.[2, 3] Potential reasons for AKI after CABG include kidney ischemia from cardiopulmonary bypass, fluctuations in blood pressure and cardiac output, systemic inflammation, oxidative stress or underlying reduced renal reserve.[4] Despite its high prevalence, it remains unclear why some patients develop post-CABG AKI and others do not, despite having similar clinical profiles and surgical courses. Current clinical prediction models focus on immediate pre- operative risk factors but lack the ability to identify individuals at high AKI risk before the immediate pre-operative period. Early identification of risk factors may provide biological insights into mechanisms leading to AKI, and may allow physicians to employ protective and preventive strategies to mitigate risk of AKI.

Conventional biomarkers of kidney health, such as estimated glomerular filtration rate (eGFR) and urine albumin-to-creatinine ratio (ACR), primarily reflect glomerular function and injury, but correlate poorly with degree of tubulointerstitial damage.[5-7] Given that kidney tubules are a major site of injury in AKI, assessment of kidney tubule health at baseline before the kidney stress from CABG may offer insights into AKI susceptibility beyond what eGFR and ACR can detect.[6] Kidney tubule biomarkers include kidney injury molecule-1 (KIM-1), a marker of tubular injury; alpha-1 microglobulin (A1M), a marker of proximal tubule resorptive capacity; and uromodulin (UMOD) and epidermal growth factor (EGF), which reflect tubule protein synthetic capacity. These biomarkers have shown promise in quantifying susceptibility to poor kidney outcomes in the ambulatory setting such as CKD progression and cardiovascular disease (CVD) events, but have not been evaluated for future risk of AKI after CABG surgery beyond the immediate pre- operative setting.[5, 8-10]

We sought to determine the associations between kidney tubule dysfunction and injury markers with future risk of AKI following CABG surgery.

## Materials and Methods

### Study Design and Data Collection – The REGARDS Cohort

The parent REasons for Geographic and Racial Disparities in Stroke (REGARDS) study and this ancillary study were approved by the Institutional Review Board of the University of Alabama at Birmingham. The parent REGARDS study obtained written consent from all cohort participants, including permission for review of medical records. The IRB waived requirement for informed consent for the ancillary study.

REGARDS is a longitudinal, population-based cohort including 30,239 community-living adults aged ≥45 years from across the continental US. Enrollment occurred between January 2003 and October 2007, and included a baseline interview where participants answered questions about demographics, health behaviors, diagnosed medical conditions and medication usage. As a part of an in-home visit, trained personnel collected vital sign measurements and obtained blood and urine samples from participants.[11] REGARDS conducted follow-up contacts with each participant at 6-month intervals for up to 19 years, identifying interim health and hospitalization events.

For this ancillary study, trained research personnel conducted structured chart reviews of hospitalizations for CABG. Medical records included Emergency Department and admission notes, surgical notes, discharge summaries, and clinical laboratory values. Abstracted information pertinent to this study included history of maintenance dialysis, kidney transplant, use of dialysis during hospitalization and all creatinine values throughout the admission.[12, 13] Research personnel had access to identifiable medical record information. The research team reviewed medical records during the period October 1, 2022 to May 1, 2024.

### Outcomes – Identification of CABG and Acute Kidney Injury

For the present study, we included participants who experienced hospitalizations associated with CABG surgery after their REGARDS baseline visit. The parent REGARDS study identified all hospitalizations and obtained copies of medical records for suspected coronary heart disease (CHD) events, which included CABG surgeries.[11, 14] For participants who had multiple CABG procedures during study follow-up, we selected the first surgery. We excluded participants who experienced a myocardial infarction or acute heart failure exacerbation during the CABG-associated hospitalization.

The primary outcome of the study was the development of AKI in the post- operative period. We defined the participant’s baseline creatinine as the lowest serum creatinine value in the 48 hours prior to CABG surgery and the post-CABG creatinine as the highest creatinine value following CABG surgery. We used the Kidney Disease Improving Global Outcomes (KDIGO) criteria for AKI, defined as a minimum rise in serum creatinine (SCr) of ≥0.3 mg/dL between the highest and baseline creatinine values.[15]

### Biomarker Analysis

Blood and urine samples obtained during the initial in-home visit were centrifuged within 2 hours of collection, placed on ice and shipped overnight to the University of Vermont. Upon arrival, laboratory researchers centrifuged the samples at 30,000 *g* and 4 °C. The biomarker measurements for the current study were performed by Brigham and Women’s Hospital. With the exception of A1M, we measured all biomarkers twice and averaged the results to improve precision. KIM- 1 and EGF were measured using a microbead based ELISA assay from Bio-Rad Bio-Plex Luminex 200 reader (Hercules, California, USA). The lower limit of detection (LLOD) and inter-assay coefficients of variation (CV) for KIM-1 was 1.98 pg/mL and 7.8%, respectively. For EGF the LLOD was 0.313 pg/mL, and the CV was 9.2%. We measured urine UMOD using the R-Plex Protocol on the Meso Scale Discovery (MSD) platform (Rockville, Maryland, USA) with a LLOD of 244.14 pg/mL and inter-assay CV of 6.4%. Lastly, we measured A1M using the SIEMENS BNII Nephelometer (Tarrytown, New York, USA) with a LLOD of 5.63 mg/L and intra-assay CV of 1.2-4.0%. This analyte was measured only once because of its low CV.

### Covariates

Covariates included demographic (age, sex, race), urine creatinine (to account for urine tonicity at time of collection), body mass index (BMI), chronic medical conditions (hypertension and diabetes mellitus) and baseline glomerular kidney measures (eGFR and urine albumin) all measured at the REGARDS baseline visit. These covariates were selected for their biologic plausibility. We identified each chronic medical condition either through the participant’s self-report or their use of disease-specific medications. Additionally, we included a diagnosis of diabetes mellitus if the participant had baseline lab values of either a fasting blood glucose of >126 mg/dL or a non-fasting blood glucose of >200 mg/dL. We calculated baseline eGFR using the participant’s baseline serum creatinine and serum cystatin C measurements according to the 2021 CKD-EPI equation without race.[16]

### Data Analysis

We reported continuous variables using means and standard deviations or medians with quartiles, as appropriate. We examined the inter-relations between biomarkers using Spearman coefficients. We performed unconditional logistic regression (univariable and multivariable) to estimate the association between each biomarker (log_2_ transformed and quartiles) with odds of AKI. The log_2_ transformation accounts for data skew, allows interpretation as “per 2-fold higher”, and facilitates comparison of strengths of associations with our prior work. We examined sequential multivariable models. Model 1 adjusted for age, sex, race, time from REGARDS interview to CABG hospitalization and urine creatinine. Model 2 included Model 1 and history of diabetes, hypertension, and BMI at REGARDS enrollment. Finally, Model 3 included Model 2 variables and eGFR and urine albumin at enrollment. Statistical significance was set at 0.05 (two-tailed). We performed all analyses using SAS version 9.4 (SAS Institute Inc., Cary, NC, USA).

## Results

Among the 30,329 REGARDS participants, 760 (2.5%) underwent CABG surgery. [17] We excluded 366 participants with illegible or incomplete records, 3 participants for a history of hemodialysis use or kidney transplant at hospital admission, 15 participants for missing serum creatinine values and 2 participants who underwent a cardiac procedure other than CABG, resulting in a final analytic sample of 394 individuals. (Figure 1) Of the 394 participants, 176 (45%) experienced AKI after their CABG surgery. Individuals who were male or Black were more likely to experience AKI. The AKI group had slightly lower eGFR values and higher urine ACR values at REGARDS baseline compared to the non-AKI group. (Table 1)

**Fig 1.**
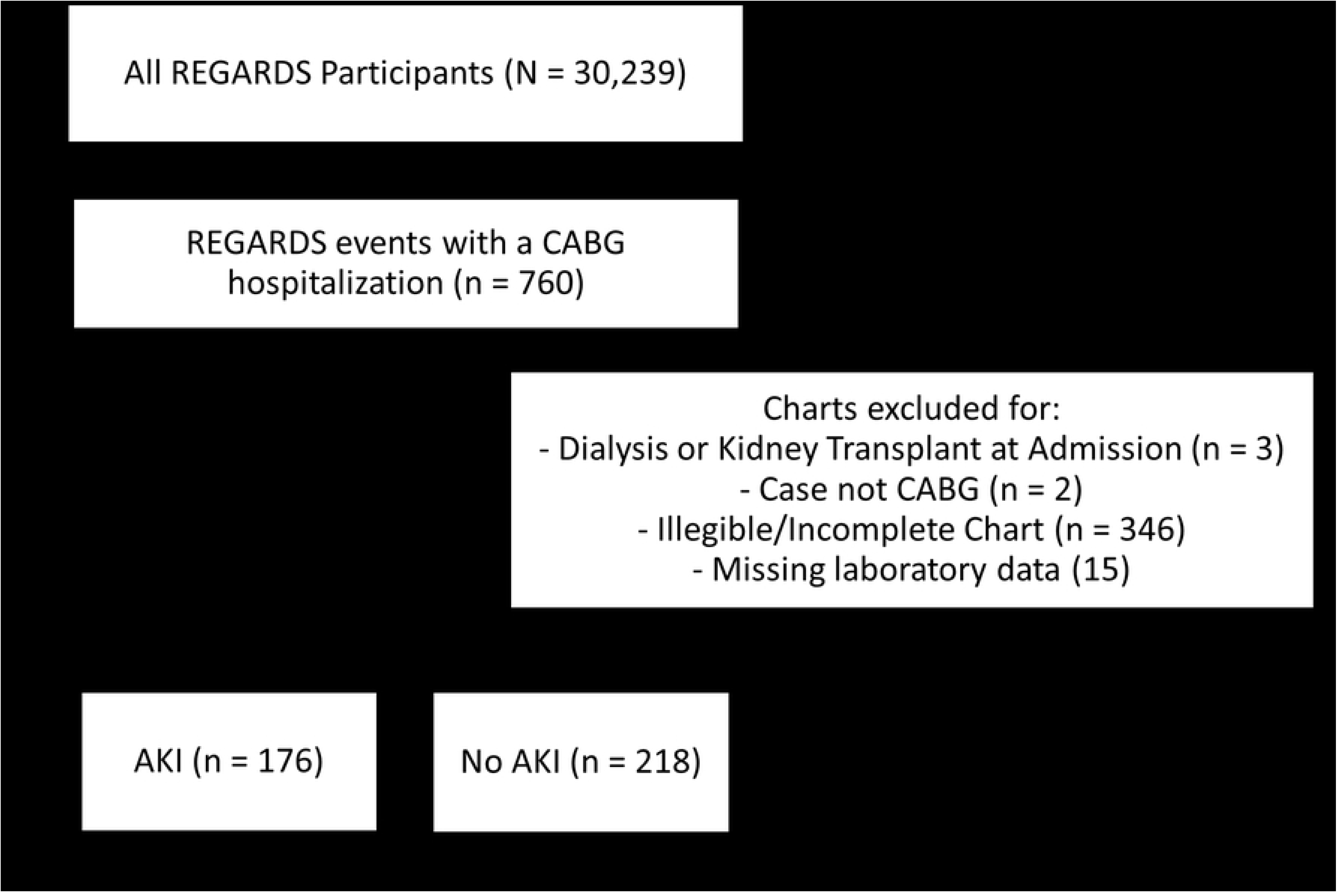
Study population. Sampling for this study within the population of participants admitted for CABG surgery in the REGARDS Trial. Abbreviations: AKI, acute kidney injury; CABG, coronary artery bypass graft; REGARDS, REasons for Geographic and Regional Differences in Stroke.

**Table 1.** Baseline Characteristics of Participants at Time of REGARDS Enrollment.

| Characteristic | AKI<br>(n = 176) | No AKI<br>(n = 218) | Total<br>(n = 394) |
| --- | --- | --- | --- |
| **Demographics** |  |  |  |
| Age, years – mean (SD) | 67 (7) | 65 (7) | 66 (7) |
| Sex, female – n (%) | 43 (24%) | 70 (32%) | 113 (29%) |
| Race, black – n (%) | 44 (25%) | 37 (17%) | 81 (20%) |
| **Health Behaviors** |  |  |  |
| Body Mass Index, kg/m<sup>2</sup> – mean (SD) | 29 (5) | 29 (5) | 29 (5) |
| **Chronic Medical Conditions** |  |  |  |
| Hypertension – n (%) | 123 (69%) | 126 (58%) | 249 (63%) |
| Diabetes – n (%) | 71 (40%) | 61 (28%) | 132 (33%) |
| **Baseline Biomarkers** |  |  |  |
| eGFR, ml/min/1.73 m<sup>2</sup> – mean (SD) | 78 (20) | 84 (17) | 81 (18) |
| Urine ACR mg/g – median (Q1, Q3) | 8.9 (5.3 – 24.6) | 7.6 (4.5 – 17.2) | 8.3 (4.8 – 20.3) |

Abbreviations: ACR, albumin to creatinine Ratio; AKI, acute kidney injury; CABG, coronary artery bypass graft; eGFR, estimated glomerular filtration rate; SD, standard deviation; Q1, first quartile; Q3, third quartile

Tubule marker values are summarized in Table 2. Urine A1M values were higher in AKI than non-AKI participants. (Table 2) Conversely, urine UMOD and EGF values were lower in AKI than non-AKI participants. KIM-1 were nearly identical in AKI vs. non-AKI participants. (Table 2) When stratified by quartiles, higher A1M was associated with lower odds of AKI, while higher urine UMOD and EGF quartiles were associated with lower odds of AKI. Higher quartiles of KIM-1 were not associated with AKI development. (Figure 2)

**Fig 2.**
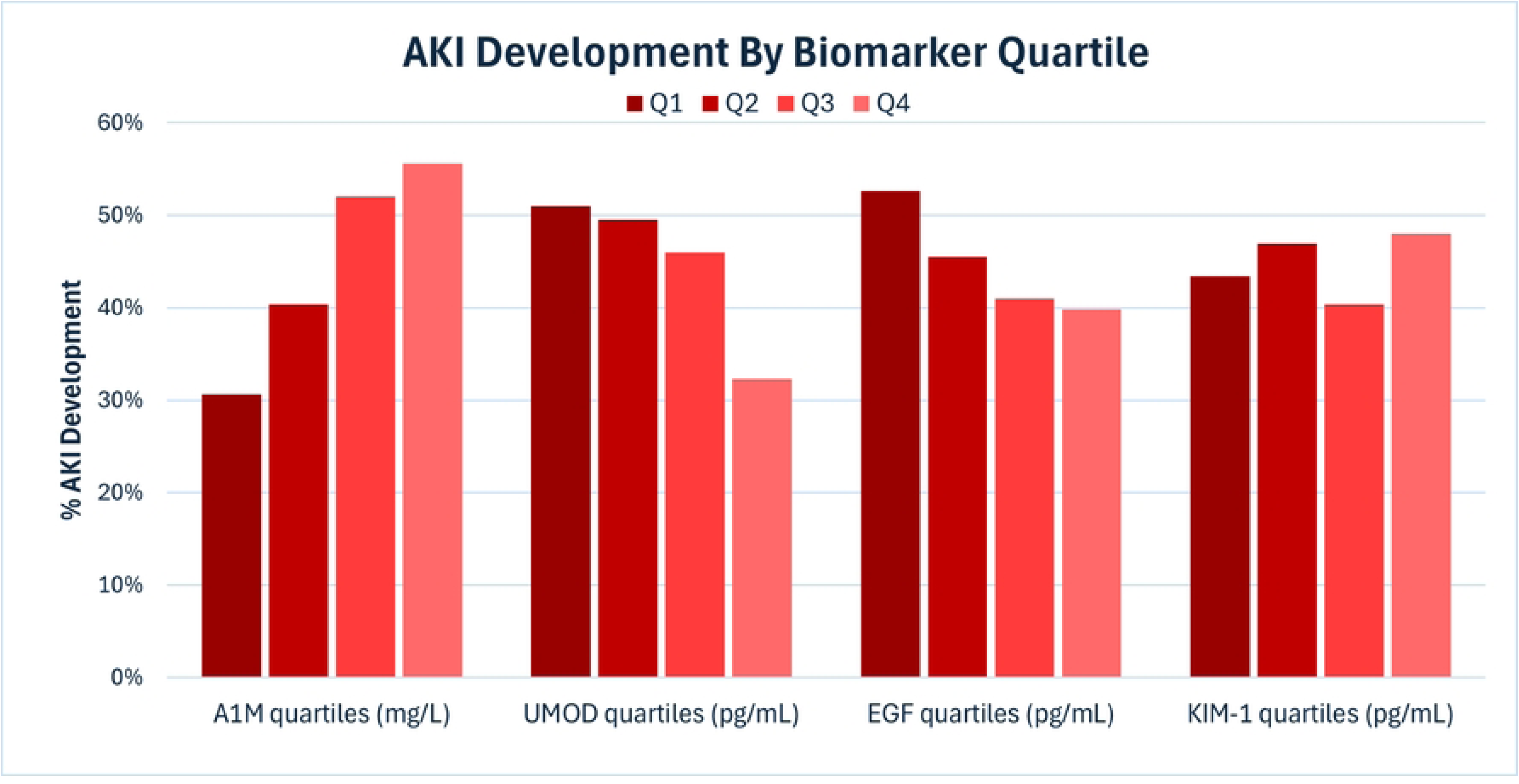
AKI Development Stratified by Biomarker Measurement Quartiles. Abbreviations: AKI, acute kidney injury; A1M, alpha-1 microglobulin; EGF, epidermal growth factor; KIM-1, kidney injury molecule-1; Q1-Q4, first through fourth quartile; UMOD, uromodulin

**Table 2.** Tubule Injury and Dysfunction Biomarker Raw Values Stratified by CABG-Associated AKI.

| Biomarkers | AKI<br>Median (Q1, Q3) | No AKI<br>Median (Q1, Q3) | Total<br>Median (Q1, Q3) |
| --- | --- | --- | --- |
| **A1M, mg/L** | 10.6 (6.3 – 16.9) | 7.8 (5.6 – 12.8) | 8.7 (5.7 – 15.1) |
| **UMOD, µg/mL\*** | 8.5 (5.2 – 13.1) | 11.3 (6.6– 17.4) | 10.0 (5.6– 15.6) |
| **EGF, ng/mL\*** | 6.6 (4.0 – 1.1) | 7.8 (4.9 – 12.5) | 7.3 (4.4 – 12.1) |
| **KIM-1, ng/mL\*** | 1.0 (0.5 – 2.1) | 1.1 (0.5 – 1.8) | 1.1 (0.5 – 1.9) |

\*Standard units are in pg/mL
Abbreviations: AKI, acute kidney injury; A1M, alpha-1 microglobulin; CABG, coronary artery bypass graft; EGF, epidermal growth factor; KIM-1, kidney injury molecule-1; Q1, first quartile; Q3, third quartile; UMOD, uromodulin

The kidney tubule markers showed weak to moderate correlation (Spearman correlation coefficient -0.12 to 0.50). (Table 3) The highest correlation (0.50) was between KIM-1 and EGF. Higher A1M was inversely correlated with eGFR. EGF and UMOD were directly correlated with eGFR, but the correlations were weak. Higher A1M was directly correlated with albuminuria, whereas EGFR and urine UMOD were inversely correlated with albuminuria.

**Table 3.** Spearman Correlation Coefficients between tubule injury and dysfunction biomarkers.

|  | **A1M/UCr** | **UMOD/UCr** | **EGF/UCr** | **KIM-1/UCr** | **eGFR** | **ACR** |
| --- | --- | --- | --- | --- | --- | --- |
| **A1M/UCr** | 1.0 | -0.07 | 0.05 | 0.26 | -0.12 | 0.25 |
| **UMOD/UCr** |  | 1.0 | -0.004 | -0.02 | 0.05 | -0.07 |
| **EGF/UCr** |  |  | 1.0 | 0.50 | 0.25 | -0.05 |
| **KIM-1/UCr** |  |  |  | 1.0 | -0.01 | 0.10 |
| **eGFR** |  |  |  |  | 1.0 | -0.01 |
| **ACR** |  |  |  |  |  | 1.0 |

Abbreviations: A1M, alpha-1 microglobulin; UMOD, uromodulin; EGF, epidermal growth factor; KIM-1, kidney injury molecule-1; UCr, urine creatinine; eGFR, estimated glomerular filtration rate; ACR, albumin:creatinine ratio

The median (interquartile range) time between REGARDS enrollment and CABG hospitalization was 5.5 years (2.7, 9.3) and did not differ with AKI stratification (AKI: 5.7 years [2.8, 9.2]; no AKI: 5.5 years [2.7, 9.4]). Median sCr baseline before CABG surgery was similar in those with (median 1.0 [0.8, 1.2]) compared to those without AKI (median 0.9 [0.8, 1.1]). Following surgery, those with AKI had larger changes in sCr (median 0.4 [0.4, 0.7]) than those who did not (median 5.5 [4, 8]) and 0.1 [0.0, 0.2], respectively), as per design.

Higher A1M was associated with higher odds of AKI following CABG (adjusted OR per 2-fold higher A1M 1.34, 95% confidence interval (CI): 1.00 – 1.80). (Table 4) This association persisted with multivariable adjustments for potential confounders, including baseline eGFR and urine albumin. Higher quartiles of A1M were similarly associated with higher adjusted odds of AKI, and the relationship appeared to increase in magnitude across quartiles. The odds of post-CABG AKI were lower with higher values of urine UMOD and EGF, albeit the association of EGF with post-CABG AKI risk did not reach statistical significance in the final model that adjusted for eGFR and albuminuria (adjusted OR per 2-fold higher urine UMOD 0.77, 95% CI: 0.62 – 0.95 and adjusted OR per 2-fold higher EGF 0.79, 95% CI: 0.59 – 1.05). When characterized by urine UMOD and EGF quartiles, the associations appeared fairly linear. We did not detect an association between urine KIM-1 and AKI following CABG (adjusted OR per each 2-fold higher KIM-1 0.92, 95% CI 0.77 – 1.10), and this finding appeared similar when evaluating KIM-1 quartiles.

**Table 4:** Associations between Biomarkers of Tubule Injury and Dysfunction and post-CABG AKI.

|  | **Unadjusted**<br>**OR (95% CI)** | **Model 1**<br>**OR (95% CI)** | **Model 2**<br>**OR (95% CI)** | **Model 3**<br>**OR (95% CI)** |
| --- | --- | --- | --- | --- |
| **<u>A1M mg/L</u>** |  |  |  |  |
| Log<sub>2</sub>(A1M) | 1.51 (1.20 – 1.89) | 1.49 (1.14 – 1.88) | 1.42 (1.09 – 1.84) | 1.34 (1.00 – 1.80) |
| Quartiles |  |  |  |  |
| Q1 | 1.00 | 1.00 | 1.00 | 1.00 |
| Q2 | 1.54 (0.85 – 2.77) | 1.50 (0.81 – 2.76) | 1.45 (0.78 – 2.70) | 1.34 (0.71 – 2.56) |
| Q3 | 2.46 (1.37 – 4.41) | 2.38 (1.26 – 4.52) | 2.24 (1.17 – 4.30) | 2.19 (1.10 – 4.36) |
| Q4 | 2.83 (1.58 – 5.08) | 2.70 (1.41 – 5.18) | 2.56 (1.32 – 4.98) | 2.24 (1.07 – 4.68) |
| **<u>UMOD, pg/mL</u>** |  |  |  |  |
| Log<sub>2</sub>(UMOD) | 0.73 (0.60 – 0.89) | 0.70 (0.57 – 0.85) | 0.72 (0.59 – 0.88) | 0.77 (0.62 – 0.95) |
| Quartiles |  |  |  |  |
| Q1 | 1.00 | 1.00 | 1.00 | 1.00 |
| Q2 | 0.94 (0.54 – 1.65) | 0.85 (0.47 – 1.51) | 0.93 (0.52 – 1.69) | 0.96 (0.52 – 1.76) |
| Q3 | 0.82 (0.47 – 1.43) | 0.75 (0.42 – 1.34) | 0.82 (0.45 – 1.50) | 0.95 (0.51 – 1.77) |
| Q4 | 0.45 (0.26 – 0.82) | 0.42 (0.23 – 0.77) | 0.48 (0.26 – 0.90) | 0.60 (0.32 – 1.15) |
| **<u>EGF, pg/mL</u>** |  |  |  |  |
| Log<sub>2</sub>(EGF) | 0.84 (0.70 – 1.01) | 0.70 (0.54 – 0.91) | 0.73 (0.56 – 0.95) | 0.79 (0.59 – 1.05) |
| Quartiles |  |  |  |  |
| Q1 | 1.00 | 1.00 | 1.00 | 1.00 |
| Q2 | 0.75 (0.43 – 1.32) | 0.59 (0.32 – 1.08) | 0.63 (0.34 – 1.17) | 0.68 (0.36 – 1.29) |
| Q3 | 0.63 (0.36 – 1.10) | 0.48 (0.26 – 0.91) | 0.51 (0.27 – 0.98) | 0.60 (0.30 – 1.20) |
| Q4 | 0.60 (0.34 – 1.05) | 0.39 (0.18 – 0.84) | 0.45 (0.21 – 0.98) | 0.61 (0.27 – 1.38) |
| **<u>KIM-1, pg/mL</u>** |  |  |  |  |
| Log<sub>2</sub>(KIM-1) | 1.04 (0.92 – 1.18) | 1.01 (0.86 – 1.19) | 0.96 (0.81 – 1.14) | 0.92 (0.77 – 1.10) |
| Quartiles |  |  |  |  |
| Q1 | 1.00 | 1.00 | 1.00 | 1.00 |
| Q2 | 1.15 (0.66 – 2.02) | 1.01 (0.58 – 1.83) | 0.94 (0.51 – 1.73) | 0.88 (0.47 – 1.65) |
| Q3 | 0.89 (0.50 – 1.55) | 0.79 (0.42 – 1.49) | 0.69 (0.36 – 1.32) | 0.66 (0.34 – 1.31) |
| Q4 | 1.20 (0.69 – 2.10) | 1.09 (0.54 – 2.21) | 0.92 (0.45 – 1.90) | 0.76 (0.35 – 1.66) |

Model 1: Adjusted for age, sex (male and female), race (black and white), time from interview to CABG (<5 and ≥5 years) and urine creatinine.
Model 2: Model 1 + adjusted for diabetes (yes and no), hypertension (yes and no) and BMI.
Model 3: Model 2 + adjusted for CKD (eGFR <60 and ≥ 60) and log<sub>2</sub>(urine albumin).
**Abbreviations:** AKI, acute kidney injury; A1M, alpha-1 microglobulin; UMOD, uromodulin; EGF, epidermal growth factor; KIM-1, kidney injury molecule-1; SCr, serum creatinine; eGFR, estimated glomerular filtration rate; OR, odds ratio; CI, confidence interval

## Discussion

Our study builds upon our prior research from the SPRINT blood pressure trial, where we found that biomarkers of kidney tubular dysfunction (A1M, UMOD, EGF), but not injury, were associated with future development of AKI in a subgroup of CKD patients.[5, 17-20] The current study suggests that markers of tubule dysfunction measured among community dwelling adults at a stable phase of health, may be associated with higher odds of AKI after future CABG surgery. These associations were independent of risk factors and glomerular kidney markers used routinely in clinical practice (eGFR and albuminuria).

Whereas most prior studies have evaluated CABG as the immediate stressor and cause of AKI, we evaluated kidney tubule health years earlier. Demirjian, et al. described a prediction model based on perioperative basic metabolic panel laboratory values with good predictive accuracy for moderate to severe acute kidney injury within 72 hours and 14 days after CABG.[21] The TRIBE-AKI cohort showed that elevated pre-surgery levels of sTNFR1, sTNFR2, and KIM-1 are associated with increased mortality, higher risk of cardiovascular events, and greater chance of CKD progression after CABG surgery, suggesting that subclinical injury and inflammation may have been present prior to the operation.[22] Our findings collectively build upon these prior studies, suggesting even earlier windows of opportunity for predicting individual vulnerability to peri-CABG AKI.

The findings of this study also provide new insights into mechanisms leading to AKI. Postulated mechanisms for AKI after CABG include patient risk factors, intraoperative hemodynamic and inflammatory insults, and postoperative complications.[23] Our findings suggest that markers of tubule dysfunction (A1M, UMOD, EGF) may be associated with future AKI risk; these markers represent proximal tubule resorption (A1M) and distal synthetic dysfunction (EGF and UMOD) and reflect the functional integrity of the kidney. Globally, these findings suggest that subtle abnormalities in kidney tubule health measured even years before the kidney stressor (CABG in this case) may indicate a reduction in the kidney’s resiliency or “renal reserve” when responding to stressors that cause AKI – changes not captured by eGFR and albuminuria. In this context, CABG may act as a type of kidney stress test where individuals with subtle and unrecognized kidney dysfunction may be more likely to develop AKI. The similarity of these findings to those of the SPRINT trial further support this hypothesis.

Markers of kidney tubule health may provide opportunities for closer monitoring, and preventive measures such as shorter bypass time, avoidance of contrast and medications such as renin-angiotensin-aldosterone axis inhibitors in the pre-operative period, and hydration procedures. We note that measures for preventing peri-CABG AKI have not yet been validated.[24] The ongoing PrevAKI Multicenter Study is evaluating whether early identification of high-risk patients using urinary biomarkers [TIMP-2]*[IGFBP7] and applying KDIGO guidelines could reduce the incidence of AKI after CABG and may offer important first insights.[25] It may be useful to add additional measures of kidney tubule dysfunction as this, and other studies, move forward.

Strengths of this study include the novel design and focus on characterizing kidney health years before AKI, as summarized above. The availability of multiple markers of kidney tubule health concurrently, and the detailed abstraction of daily creatinine values during the CABG admissions are additional strengths. The study builds off hypotheses generated in another cohort with different etiologies of AKI, but provides similar findings, strengthening confidence in the inferences. Finally, with the exception of A1M which has a very low CV, all biomarkers were measured in duplicate and averaged to improve precision. The study also had important limitations. The REGARDS cohort is limited to individuals ≥ 45 years. We retrospectively abstracted clinical data through manual review of digital images of paper records. We did not differentiate the extent, types or methods of CABG surgery, so we cannot differentiate elective from urgent surgical procedures.

Although we presume that AKI episodes following CABG were due to the procedure, other factors may have triggered AKI. We determined AKI based upon serum creatinine values available in the medical record. While we had serum creatinine values at REGARDS baseline years earlier, we did not have access to pre-hospitalization serum creatinine values prior to the CABG admission. Lastly, there is the possibility of biomarker degradation over time, between the collection and the biomarker measurements; however, this would have biased results towards the null hypothesis.

## Conclusion

Biomarkers of tubule health, measured in community dwelling adults at a stable phase of health, are associated with AKI events after future CABG surgery. This finding was stronger for markers of kidney tubule dysfunction than for markers of tubule injury. Higher A1M and lower urine UMOD were associated with higher odds of AKI, independent of eGFR, albuminuria, and AKI risk factors. These findings provide new insights into the mechanisms of AKI after bypass surgery and provide opportunities to identify higher risk patients for post-CABG AKI where preventive strategies may be warranted.

## Data Availability

Data for this study may be obtained upon request from the Reasons for Geographic and Racial Disparities in Stoke (REGARDS) study, https://www.uab.edu/soph/regardsstudy/.

## Acknowledgements

We would like to acknowledge the following individuals for contributions to medical record abstraction: Linda Cho, Ching-Min (Bryan) Chu, Derick Campbell, Eliane De Jong, Clemence Gatete, Cheryl Herchek, Emma Helkey, Emma Helwagen, Ireland Hester, Alexander Keister, Dina McGowan, Isabella Rigg, Riley Riggenbach, John Rider, and Lauren Shingler.

## Key words and abbreviations

AKI: acute kidney injury
CKD: chronic kidney disease
ESRD: end-stage renal disease
A1M: alpha-1 microglobulin
UMOD: uromodulin
EGF: epidermal growth factor
KIM-1: kidney injury molecule-1
CABG: coronary artery bypass graft
cohort study
SCr: serum creatinine

